# Machine learning-assisted serum Raman spectroscopy for risk stratification in men with suspected prostate cancer: a retrospective proof-of-concept study

**DOI:** 10.64898/2026.10.26.26366514

**Authors:** Kirsty Milligan, Ian Smith, Anton Flugge, Nuha Fakhrua Aldeen, Brian Shine, Cecilia Muldoon

## Abstract

**Background:** Serum prostate-specific antigen (PSA) remains the principal laboratory investigation for men with suspected prostate cancer but has limited specificity, particularly in patients with PSA concentrations between 4 and 10 ng/mL. Additional blood-based tests capable of improving risk stratification before imaging or biopsy could reduce unnecessary investigations while maintaining diagnostic sensitivity. Raman spectroscopy provides a biochemical fingerprint of biological samples and, when combined with machine-learning methods, may offer a novel approach to serum-based cancer detection.

**Methods:** Archived serum samples obtained from men with histologically confirmed prostate cancer and symptomatic control subjects were analysed using dried-serum Raman spectroscopy. Raman spectra were incorporated into supervised machine-learning classification models to generate predicted probabilities of prostate cancer. Diagnostic performance was evaluated in the complete cohort and separately in patients with PSA concentrations between 4 and 10 ng/mL.

**Results:** The study included 119 men with prostate cancer and 60 symptomatic controls. Predicted probabilities of prostate cancer were significantly higher in cancer cases than controls. The classifier demonstrated moderate discrimination in the complete cohort (AUC approximately 0.77) and maintained similar performance in patients with PSA concentrations between 4 and 10 ng/mL (54 prostate cancer and 41 symptomatic controls, AUC approximately 0.78).

**Conclusions:** Machine learning-assisted analysis of dried-serum Raman spectra shows promise as an adjunct to PSA testing. The maintenance of diagnostic performance within the PSA grey zone suggests a potential role as a second-line risk-stratification test before MRI or biopsy. Prospective validation within contemporary diagnostic pathways is now required.

## Introduction

Prostate cancer remains one of the commonest malignancies affecting men worldwide and is a leading cause of cancer-related mortality.^1,2^ Earlier diagnosis offers the opportunity for curative treatment but must be balanced against the risks of unnecessary investigation and over-diagnosis of indolent disease.^3,4,5,6^

Although PSA remains the cornerstone of laboratory investigation in men with suspected prostate cancer, its specificity for clinically significant disease is limited because concentrations may also be elevated in benign prostatic hyperplasia, prostatitis and other non-malignant conditions. This has led to the development of multivariable risk assessment tools that combine PSA isoforms with clinical variables, including the Prostate Health Index (PHI)^7^ and the four-kallikrein (4Kscore) panel^8^ developed by Lilja and colleagues, together with urinary molecular tests such as SelectMDx^9^ and ExoDx.^10^ While these approaches improve risk stratification, uncertainty remains, particularly in men with PSA concentrations between approximately 4 and 10 ng/mL, where decisions regarding MRI and biopsy continue to be challenging.^11^ The objective of modern prostate cancer diagnostics is not simply to detect cancer, but to identify men with clinically significant disease who are most likely to benefit from further investigation and treatment.^12^

Multiparametric magnetic resonance imaging (mpMRI) has substantially improved diagnostic pathways and is now recommended before biopsy in many healthcare systems.^11,13^ Nevertheless, MRI is relatively expensive, requires specialist expertise and remains a finite healthcare resource. Additional blood-based tests capable of refining cancer risk before MRI could improve patient selection, reduce unnecessary imaging and increase the efficiency of diagnostic pathways.^14^

Raman spectroscopy is a vibrational spectroscopic technique that provides a molecular fingerprint of biological samples without requiring targeted molecular assays. Recent advances in instrumentation and machine-learning methods have substantially increased interest in its application to clinical diagnostics. Rather than measuring individual biomarkers, Raman spectroscopy captures global biochemical information that may reflect disease-associated molecular changes in serum.^15^

Several groups have demonstrated the feasibility of Raman spectroscopy for distinguishing patients with cancer from controls.^16,17,18,15,19,20,21,22^ However, relatively few studies have evaluated how Raman-based assays might be integrated into established clinical pathways or examined their potential role alongside PSA testing.

In the present proof-of-concept study we evaluated a machine learning-assisted dried-serum Raman assay using archived serum samples from men undergoing investigation for suspected prostate cancer. Particular emphasis was placed on the clinically important subgroup with PSA concentrations between 4 and 10 ng/mL, where improved risk stratification could assist decisions regarding further investigation.

## Methods

### Study design

This retrospective proof-of-concept study evaluated the performance of a machine learning-assisted serum Raman spectroscopy assay for the discrimination of prostate cancer from non-malignant urological disease. Archived serum samples obtained from a reference biobank were analysed using a standardised dried-serum Raman spectroscopy protocol, and model-derived probabilities of prostate cancer were compared with the reference clinical diagnosis.

### Patient cohort

Serum samples were obtained from the QUAlity biobanking iNvesTigating Urological Malignancies (QUANTUM) biobank, Nuffield Department of Surgical Sciences, University of Oxford.^23^

To enable comparison with PSA-based diagnostic pathways, only participants with a serum PSA measurement obtained within two months before or two days after serum collection were included in the present analysis.

The final study cohort comprised 179 men, including 119 patients with histologically confirmed prostate cancer and 60 symptomatic control subjects without histological evidence of prostate cancer. Sixteen control subjects had atypical benign pathological findings including prostatic intraepithelial neoplasia (PIN) and/or atypical small acinar proliferation (ASAP). The patient cohort analysed for the PSA 4-10 ng/mL model included 54 confirmed prostate cancer cases and 41 symptomatic controls).

Control subjects were recruited through the same clinical pathway as cases and are therefore representative of men undergoing investigation for suspected prostate cancer rather than healthy volunteers. Clinical details for the complete cohort are described in Table 1.

**Table 1.**
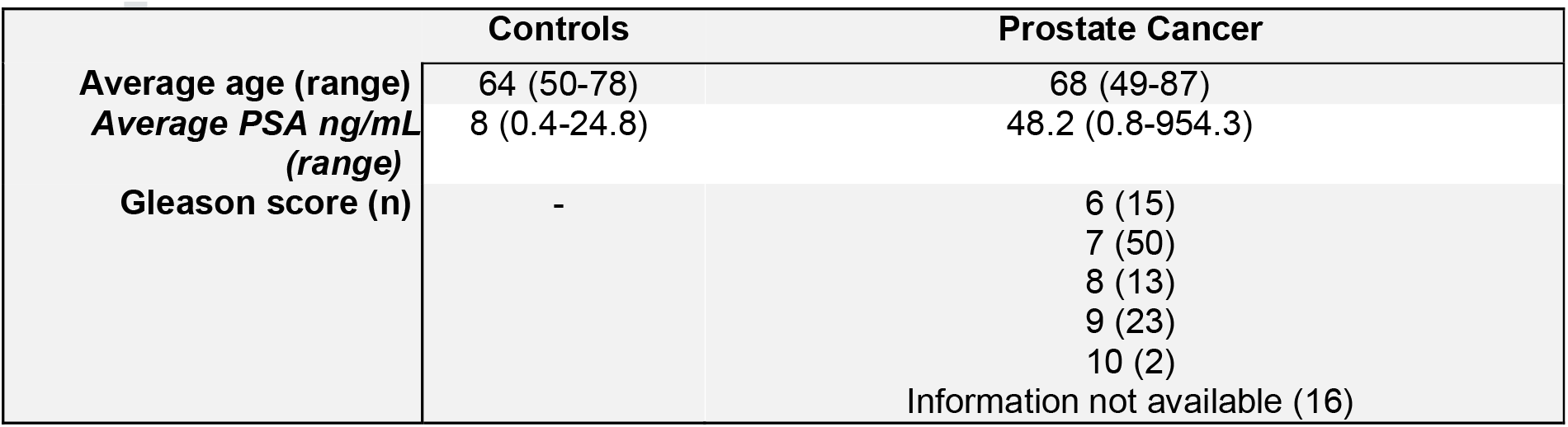
Cohort details.

|  | Controls | Prostate Cancer |
| --- | --- | --- |
| Average age (range) | 64 (50-78) | 68 (49-87) |
| Average PSA ng/mL (range) | 8 (0.4-24.8) | 48.2 (0.8-954.3) |
| Gleason score (n) | - | 6 (15)<br>7 (50)<br>8 (13)<br>9 (23)<br>10 (2)<br>Information not available (16) |

### Serum preparation

Frozen serum samples were thawed at room temperature immediately before analysis. Ten microlitres of serum were deposited onto aluminium foil mounted on a glass microscope slide and allowed to dry under ambient laboratory conditions prior to Raman spectral acquisition.

### Raman spectroscopy

Raman spectra were acquired using a Wasatch Photonics WP-785 Raman spectrometer equipped with a fibre-coupled 785 nm excitation laser. The nominal acquisition parameters are described in Table 2.

**Table 2.** Raman spectroscopy acquisition parameters.

| Parameter | Value |
| --- | --- |
| Excitation wavelength | 785 nm |
| Laser power | 450 mW |
| Acquisition time | 500 ms |
| Spot size | 120 μm |
| Spectral range | 270–2700 $\text{cm}^{-1}$ |
| Spectral resolution | 7 $\text{cm}^{-1}$ |

### Spectral preprocessing and machine learning analysis

Fifteen spectra were acquired from each dried serum spot and averaged to generate a single representative spectrum for each participant.

The signal to noise ratio of each spectrum was evaluated by computing the ratio of four defined peaks in the spectrum with noise in a silent region of the spectrum. Processed Raman spectra were incorporated into supervised machine-learning classification models to generate a predicted probability of prostate cancer for each participant. The average Raman spectrum for prostate cancer individuals and symptomatic controls, as well as tentative assignments of spectral features to molecule class, are shown in supplementary Figure S1.^24^

Separate models were evaluated for:

1. Raman spectra combined with PSA concentration and age.
2. Participants with PSA concentrations between 4 and 10 ng/mL, representing the diagnostic grey zone.

All models were based on a gradient boosting tree model (xgboost)^25^ and evaluated using five-fold cross-validation. The output of the models are probability scores representing the likelihood of that a sample belongs to the prostate cancer cohort. Principal component analysis (PCA) was employed to reduce the dimensionality of the spectra. The model was tested 100 times using different random folds each time, to ensure robustness of output. All reported results are the average of 100 repetitions of the model, unless stated otherwise.

### Statistical analysis

Predicted probabilities were summarised graphically using box plots with overlaid individual observations.

Diagnostic discrimination was evaluated using receiver operating characteristic (ROC) analysis. Area under the ROC curve (AUC), sensitivity and specificity were calculated for each model.

Separate analyses were performed for the complete cohort and for participants with PSA concentrations between 4 and 10 ng/mL.

Pre-processing and modelling were performed using Python 3.12, further analyses were performed using R version 4.6.0 and Python 3.12.

## Results

### Study cohort

A total of 179 participants were included in the analysis, comprising 119 men with histologically confirmed prostate cancer and 60 symptomatic control subjects, of which 54 cancer patients and 41 control subjects had a PSA measurement between 4 and 10 ng/mL. To permit comparison with PSA-based diagnostic pathways, only participants with serum PSA measurements obtained within two months before, or two days after, serum collection were included.

### Distribution of predicted probabilities

Figure 1 illustrates the distribution of model-derived probabilities of prostate cancer for both diagnostic groups. With the exception of the PSA + Age model in the PSA 4-10 cohort, all median predicted probabilities were higher in men with prostate cancer than in symptomatic controls, although overlap between groups was observed.

**Figure 1.**
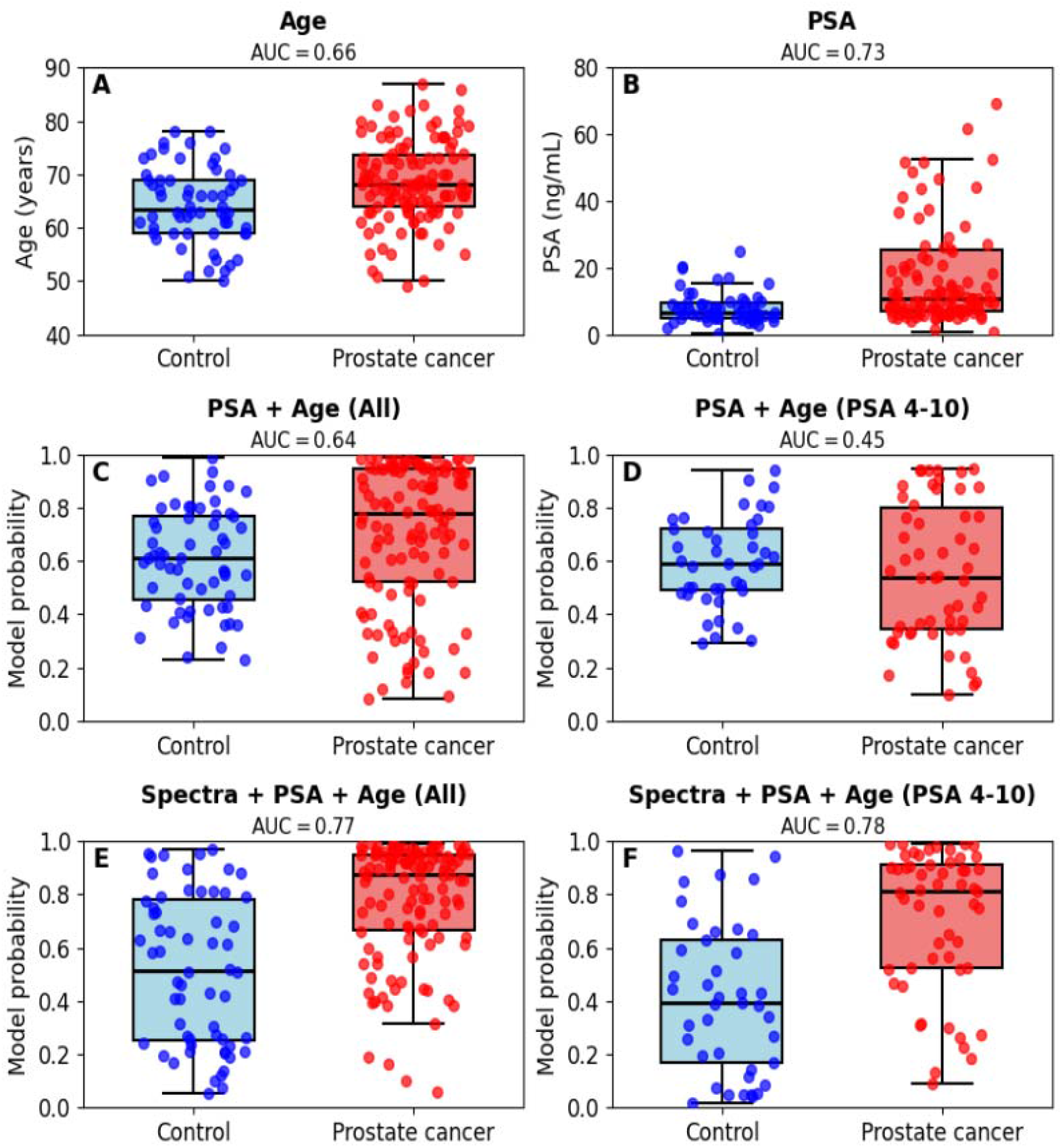
Boxplots depicting differences in distribution between control (blue) and prostate cancer (red) groups using (a) Age (b) PSA values, **(c)-(f)** the average model prediction for an individual to belong to the prostate cancer cohort for the different models; **(c)** PSA + Age for complete cohort **(d)** PSA + Age for PSA 4-10 cohort **(e)** SEROX Raman spectra + PSA + Age model for complete cohort and **(f)** SEROX Raman spectra + PSA + Age for PSA 4-10 cohort. AUC values are stated for each model.

The classifier incorporating Raman spectra, PSA concentration and age demonstrated separation between diagnostic groups in the complete cohort. Similar discrimination was observed within the subgroup of men whose PSA concentrations lay between 4 and 10 ng/mL.

### Diagnostic performance

Receiver operating characteristic (ROC) curves for the complete cohort and the PSA grey-zone subgroup are shown in Figure 2.

**Figure 2.**
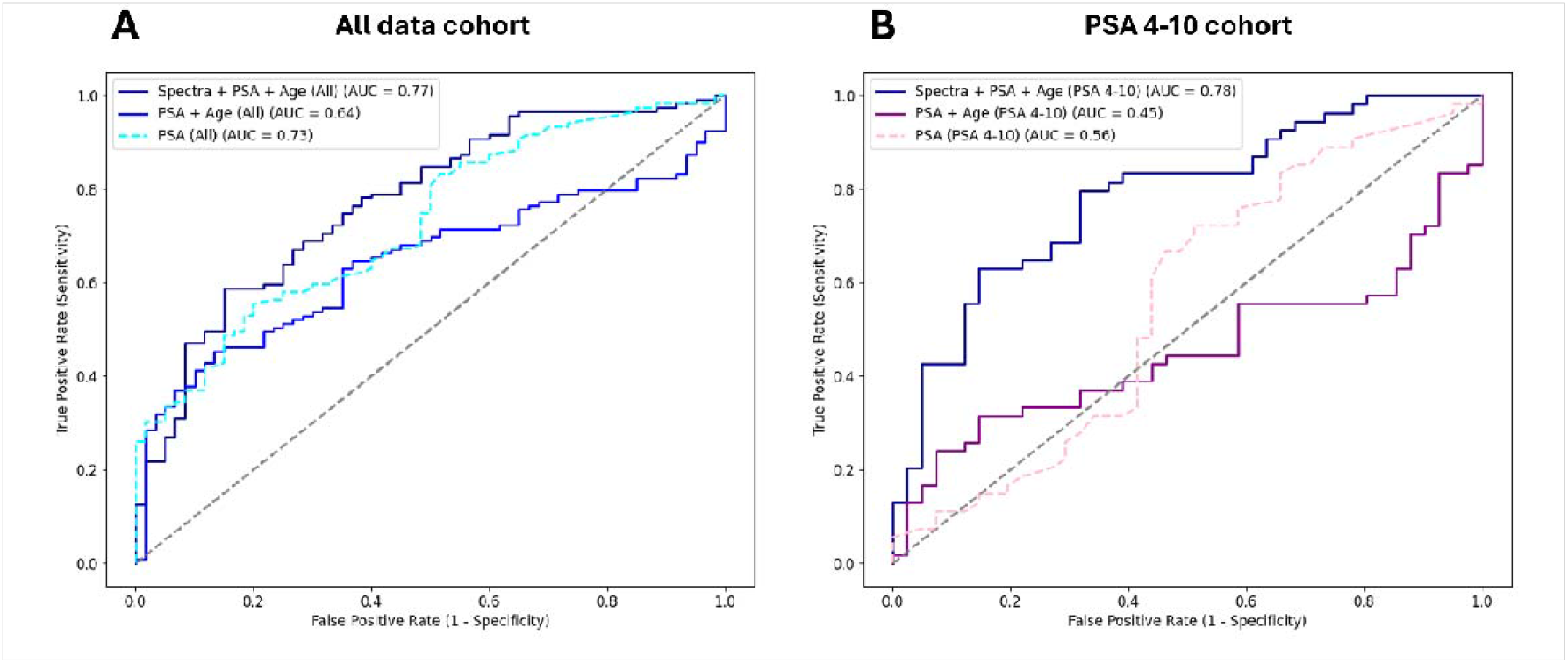
ROC curves comparing PSA, PSA + Age and SEROX Raman spectra + PSA + Age for **(a)** complete sample cohort and **(b)** PSA 4-10 cohort.

The classifier demonstrated moderate discrimination in the complete cohort, with an area under the ROC curve (AUC) of approximately 0.77. Diagnostic performance was maintained in men with PSA concentrations between 4 and 10 ng/mL, where the AUC was approximately 0.78.

The similarity in diagnostic performance between the complete cohort and the PSA grey-zone subgroup suggests that the Raman-derived molecular signature retains discriminatory capability within the clinically challenging range of PSA concentrations.

## Discussion

The present study demonstrates the feasibility of combining dried-serum Raman spectroscopy with machine-learning analysis to generate clinically meaningful estimates of prostate cancer probability. Although the overall diagnostic performance was moderate, discrimination was maintained within the clinically important PSA grey zone,^11,26^ suggesting a potential role for Raman spectroscopy as a second-line risk stratification test.

Current prostate cancer diagnostic pathways increasingly emphasise the identification of clinically significant disease while reducing unnecessary imaging and biopsy. Serum PSA remains the principal laboratory investigation, but its limited specificity has driven the development of multivariable risk assessment tools such as the Prostate Health Index (PHI), the four-kallikrein (4Kscore) panel and several urinary molecular assays.^14,7,3,9,10^ Rather than replacing these approaches, Raman spectroscopy offers a fundamentally different strategy by interrogating the global biochemical composition of serum rather than measuring predefined molecular targets.

The proposed role of the present assay is therefore as an adjunct to existing diagnostic pathways rather than as a replacement for PSA or multiparametric MRI. One potential application would be as a reflex test in men with PSA concentrations between 4 and 10 ng/mL, where uncertainty regarding the need for MRI or biopsy remains greatest,^26^ as demonstrated in Figure 3. The observation that diagnostic performance was maintained within this subgroup supports this proposed clinical role, although prospective validation in contemporary diagnostic pathways will be required.

**Figure 3.**
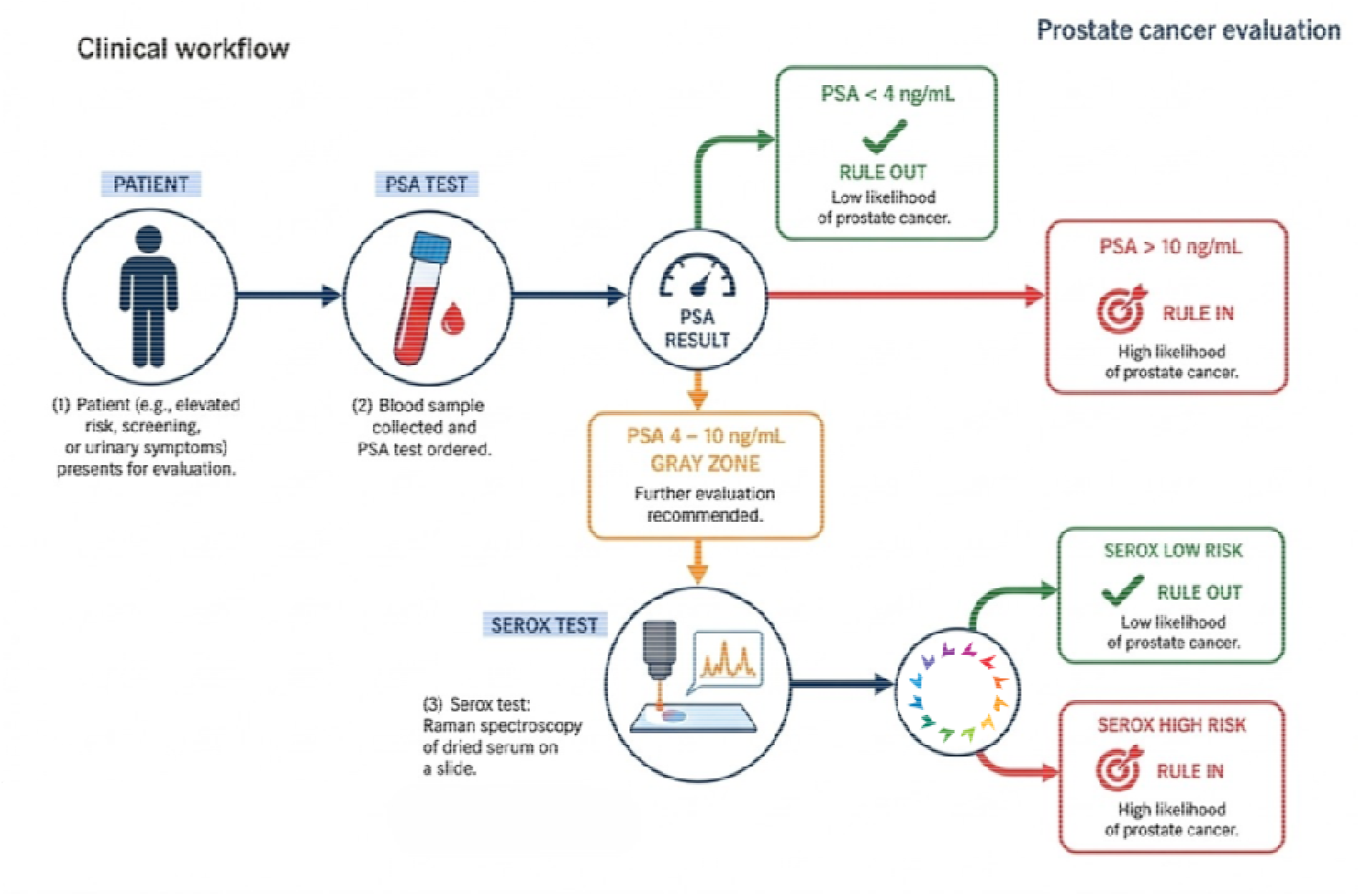
Proposed clinical utility of SEROX Raman test as a reflex test for individuals within the PSA “grey-zone” of 4-10ng/mL.

The control cohort used in the present study comprised symptomatic men undergoing investigation for suspected prostate cancer rather than healthy volunteers. Sixteen controls had atypical pathological findings including PIN and/or ASAP. Although these individuals were classified as controls for the purposes of model development, both lesions are recognised markers of increased future prostate cancer risk. Consequently, some apparent false-positive classifications may represent underlying biological changes not captured by the reference diagnosis available at the time of serum collection.

The principal strengths of this study are the use of clinically relevant symptomatic controls and the evaluation of the classifier within the PSA grey zone, where improved risk stratification is most likely to influence patient management. The use of serum also offers practical advantages because blood sampling is already embedded within established diagnostic pathways and may prove easier to standardise than urine-based assays.

Several limitations should also be acknowledged. This was a retrospective proof-of-concept study using archived samples from a single biobank, and external validation has not yet been performed. The machine-learning classifier requires prospective evaluation in independent cohorts’ representative of current clinical practice, particularly in pathways incorporating routine multiparametric MRI. Additional work will also be required to determine whether Raman-derived molecular signatures can distinguish clinically significant prostate cancer from indolent disease, which represents the principal objective of contemporary prostate cancer diagnostics.

In conclusion, machine learning-assisted serum Raman spectroscopy represents a promising approach to molecular risk stratification in men undergoing investigation for suspected prostate cancer. Although further technical and clinical validation is required, maintenance of diagnostic performance within the PSA grey zone suggests that this approach merits prospective evaluation as an adjunct to existing diagnostic pathways.

## Data Availability

This research has been conducted with support from the QUANTUM (QUAlity biobanking iNvesTigating Urological Malignancies) Research Tissue Bank, which is operated by the University of Oxford.

## Declarations

## Author contributions

Conceptualisation: CM, KM, IS, AF, NFA. Methodology: KM, NFA, AF. Machine learning analysis: AF, BS. Writing: IS, KM, BS. Writing, reviewing and editing: All authors.

## Ethics approval and consent to participate

This study was performed under the ethics approval granted by North of Scotland Research Ethics Service to QUANTUM Research Tissue Bank on 15 February 2022 (REC reference 22/NS/0004, IRAS project ID: 228721). The Committee confirmed that the favourable ethical opinion applies to all research projects conducted in the UK using tissue or data supplied by the tissue bank, provided that the release of tissue or data complies with the stated conditions; and that it is not be necessary for these researchers to make project-based applications for ethical approval, as they will be deemed to have ethical approval from this committee.

Experiments were performed in accordance with the World Medical Association Declaration of Helsinki in a facility equipped to Biosafety Level 2. The laboratory has an HTA licence (12811) and is ISO 20387:2018 (Biobanking) accredited (31342).

## Availability of data and materials

The processed model outputs underlying Figures 1 and 2 (per-sample mean predicted probabilities with class labels and age) may be made available from the corresponding author on reasonable request, subject to approval by the QUANTUM Research Tissue Bank and a data-sharing agreement.

## Consent for publication

Not applicable

