## Supplemental Figure 1 for "Machine learning-assisted serum Raman spectroscopy for risk stratification in men with suspected prostate cancer: a retrospective proof-of-concept study"

### Supplementary Information

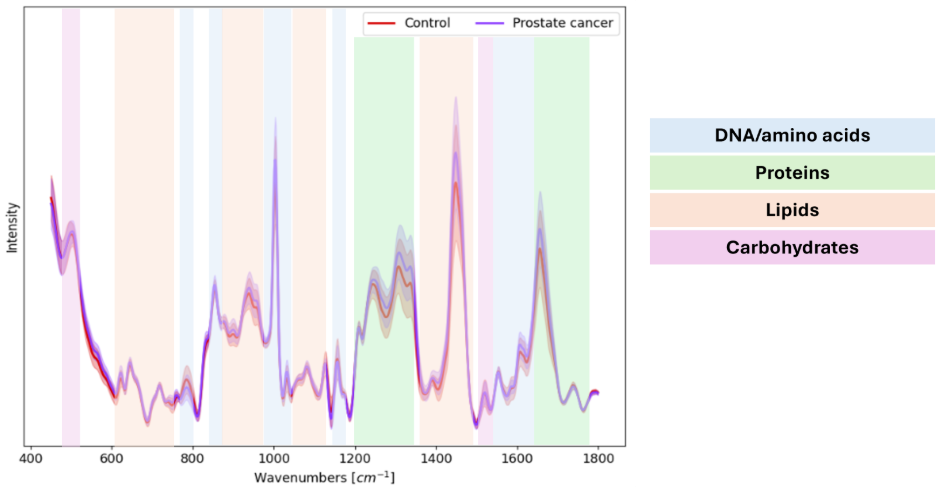

**Figure S1** Average Raman spectrum of dried serum acquired from prostate cancer positive (purple) and symptomatic control (red) individuals, after spectral processing. Shaded spectrum represents ± 1 standard deviation. Spectral features have been tentatively assigned to four major classes of biological components; DNA/amino acids (blue), proteins (green), lipids (orange), and carbohydrates (purple).
